# COVID-19 in Latin America: Contrasting phylodynamic inference with epidemiological surveillance. (Molecular epidemiology of COVID-19 in Latin America)

**DOI:** 10.1101/2020.05.23.20111443

**Authors:** Diana M. Rojas-Gallardo, Sandra C. Garzón-Castaño, Natalia Millán, Erika V. Jiménez-Posada, Marlen Martínez-Gutiérrez, Julian Ruiz-Saenz, Jaime A. Cardona-Ospina

**Affiliations:** Grupo de Investigación Biomedicina, Facultad de Medicina, Fundación Universitaria Autónoma de las Américas, Pereira, Risaralda, Colombia; Semillero de Investigación en Infecciones Emergentes y Medicina Tropical, SIEM, Fundación Universitaria Autónoma de las Américas, Pereira, Risaralda, Colombia; Public Health and Infection Research Group, Faculty of Health Sciences, Universidad Tecnológica de Pereira, Pereira, Risaralda, Colombia; Emerging Infectious Diseases and Tropical Medicine Research Group. Instituto para la Investigación en Ciencias Biomédicas - Sci-Help, Pereira, Risaralda, Colombia; Grupo de Investigación en Ciencias Animales - GRICA, Facultad de Medicina Veterinaria y Zootecnia, Universidad Cooperativa de Colombia, Bucaramanga, Colombia; Infettare, Facultad de Medicina, Universidad Cooperativa de Colombia, Medellín, Colombia

**Keywords:** SARS-CoV-2, COVID-19, Phylodynamic, Molecular Epidemiology, Latin America

## Abstract

**Background:** SARS-CoV-2 revealed important gaps in infectious disease surveillance. Molecular epidemiology can help monitoring and adapting traditional surveillance to surpass those limitations. This work aims to contrast data driven from traditional surveillance with parameters inferred from molecular epidemiology in Latin America (LATAM)

**Methods:** We obtained epidemiological data up to 4^th^ June, 2020. We estimated Effective Reproductive Number (Re) and epidemic curves using maximum likelihood (ML). SARS-CoV-2 genomes were obtained from GISAID up to June 4^th^ 2020. We aligned sequences, generated a ML phylogenetic tree, and ran a coalescent model Birth Death SIR. Phylodynamic analysis was performed for inferring Re, number of infections and date of introduction.

**Findings:** A total of 1,144,077 cases were reported up to 4^th^ June 2020. Countries with the largest cumulative cases were Chile, Peru and Panama. We found at least 18 different lineages circulating, with a predominance of B.1 and B.1.1. We inferred an underestimation of the daily incident cases. When contrasting observed and inferred Re, we did not find statistically significant differences except for Chile and Mexico. Temporal analysis of the introduction of SARS-CoV-2 suggested a detection lag of at least 21 days.

**Interpretation:** Our results support that epidemiological and genomic surveillance are two complementary approaches. Even with a low number of genomes proper estimations of Re could be performed. We suggest that countries, especially developing countries, should consider to add genomic surveillance to their systems for monitoring and adapting epidemiological control of SARS-CoV-2.

## Introduction

SARS-CoV-2 (Severe Acute Respiratory Syndrome Coronavirus - 2) the causative agent of COVID-19, has revealed the need for better, more accurate, more expedite, and widely available surveillance tools, for improving infectious disease detection and control. The fact that COVID-19 control relies up to this moment on proper disease surveillance, due to the lack of an approved vaccine or effective treatment, obliged to promptly establish early detection strategies of cases and contacts for rapid implementation of patient isolation.^1^ However, traditional epidemiological surveillance methods are subjected to limitations inherent to the ability of each health system of capturing and properly diagnosing and following symptomatic and asymptomatic cases and contacts.^2^

These limitations lead often to underestimation of the number of cases and real burden of the disease,^3^ and are usually overcome through community based studies, like for example serological surveys. These, allow an “a posteriori” estimation of the expansion factor for adjusting the real number of cases during an epidemic.^4^ Notwithstanding, in the context of the current pandemic, there is a need for almost real-time assessment of case underestimation for adjusting containment measures and facilitate disease control. In Latin American countries (LATAM) this is particularly important since differences in demography, social structure and health care availability can limit the effect of strategies for controlling the spread of the virus.^5^.

In this context, as new molecular tools have been integrated into disease surveillance, and mathematical models for the understanding of the relationship of pathogen evolution and transmission dynamics have been developed,^6,7^ molecular epidemiology has presented solutions to the limitations of traditional surveillance systems.^8^ Phylodynamic inference allows the estimation of susceptible, infected and recovered cases, and phylogeographic analysis allows the assessment of transmission chains and temporospatial distribution of infections.^7^ Therefore, this study pretends to contrast data driven from epidemiologic surveillance in LATAM, after the first one hundred days of pandemic, with parameters inferred from phylodynamic analysis of reported genomes of SARS-CoV-2 across different countries, in order to compare the inferred versus the observed number of cases, and perform a temporal analysis of the introduction of SARS-CoV-2 to each country.

## Materials and Methods

### Study design and setting

This is a molecular epidemiology study based on public epidemiologic and genomic data of SARS-CoV-2 in LATAM up to 4^th^ June, 2020 (One hundred days after the first detection of the virus in Brazil on February 25, 2020)^9^. For its reporting, we adhered the Strengthening the Reporting of Molecular Epidemiology for Infectious Diseases (STROME-ID) statement.^10^

### Data sources

COVID-19 epidemiological surveillance data was obtained from data published by the European Centre for Disease Prevention and Control until 4^th^ June, 2020, including only countries belonging to LATAM as defined by the Pan American Health Organization (PAHO). Epidemiological information obtained from this surveillance was contrasted with information inferred from phylodynamic analysis of complete genomes of SARS-CoV-2 in LATAM reported to GISAID up to 4^th^ June, 2020.^11^ Information about laboratories that reported this information is publicly available at https://www.gisaid.org/. We searched for sequences reported for North and South America up to June 4^th^ 2020. We choose only whole genome sequences (>29,900) with high coverage, and more than 95% of nucleotide base resolved belonging to countries from LATAM as previously defined.

### Epidemiological analysis

Daily incident cases, dates of introduction and fatal cases were obtained. We calculated Case Fatality Rates (CFR) = number of reported deaths/number of total cases) × 100, and estimated the effective reproductive number (Re) and epidemiologic curves using maximum likelihood (ML) methods with the *R0 v 1*.*2-6* package for R ^12,13^. In order to estimate Re, we estimated the distribution of the generation time (GT), considered as the time between a primary infected case and a secondary case, through ML using the serial time between infectee/infector pairs published by He *et al*, 2020.^12,14^ Then, Re at the beginning of the epidemic, and its evolution through time were estimated.^13^ The time period over which growth was exponential was estimated using the deviance R-squared statistic over a range of possible time periods.^12^

### Phylogenetic analysis

We conserved identical sequences with different sampling dates and different location and discarded identical sequences with same sampling date within countries. Accession IDs, sampling dates and locations of the sequences were obtained. We also generated subgroup datasets for each country reporting at least five sequences for performing phylodynamic analysis. Sequences were aligned using MAFFT software v7.450 and visually inspected and edited using Bioedit v7.0.5.3. To assess for viral recombination, we employed the pairwise homoplasy index (PHI) test using SplitsTree v4.15.1. Nucleotide substitution model for the alignment was inferred according to the corrected Akaike information criterion (AICc) in jModelTest v2.1.10. The ML tree was inferred using PhyML software V3.0 (DOI: 10.1093/sysbio/syq010) with an approximate likelihood-ratio test (aLRT) approach. Phylogenetic lineages were defined as previously reported by Rambaut et al.,^15^ by using the PANGOLIN (Phylogenetic Assignment of Named Global Outbreak LINeages) COVID-19 Lineage Assigner web tool (Available at https://pangolin.cog-uk.io/).

### Coalescent and Phylodynamic analysis

In order to infer and contrast molecular epidemiology data about cumulative infections and Re, we performed a phylodynamic analysis for each country with at least five included sequences. For each country data set, coalescent model Birth Death SIR (serial) ^16^ was run with *Phylodynamics* package in BEAST v2.6.2.0. Priors were introduced with BEAUti and all data sets were run with the same prior distribution and value for Molecular clock rate (0.0006), Become Uninfectious Rate (Normal (mean=37.0, sigma =1.3)) and Reproductive Number (Log-normal (mean log=0,0, sd log =1.2)). Markov chain Monte Carlo (MCMC) runs of 80 million states sampling every 8,000 steps were computed. The convergence of MCMC chains was checked using Tracer v.1.7.1, ensuring that the effective sample size (ESS) values were greater than 300 for each parameter estimated; for each data set, the maximum clade credibility (MCC) tree was obtained from the trees posterior distribution using TreeAnnotator after 10% burn-in. Finally, time scaled phylogenies, substitution rates, and TMRCA were estimated using the *treedater* R package 0.5.0 assuming an strict molecular clock. Re evolution through time was estimated using the *skyspline* R package 0.1, and SIR trajectories were calculated using the *phydynR* R package 0.2.0^17^ using an exponential growth model using birth rates, death rates and effective population number (Ne) estimated by BEAST for each country.

### Sensitivity analysis

We conducted a sensitivity analysis computing the R-squared statistic over a range of possible time periods for exponential growth (begin between days 1 to 50, and ending between days 51 to 100), selecting the best fit for ML analysis. In order to assess the effect of GT distribution variation on Re estimation, we computed the reproduction ratio over a range of chosen GT distributions with mean between 1 to 14 days, assuming fixed standard deviation, and using ML method.

### Role of the funding source

We did not have any funding source for funding this research. All the authors had full access to all the data in the study and had final responsibility for the decision to submit for publication.

## Results

### Epidemiological analysis of COVID-19 in LATAM

A total of 1,144,077 cases were reported up to 4^th^ June 2020, with 57,453 deaths (overall CFR 3·45%). At the day 100 after the introduction of COVID-19 in LATAM, the countries with largest number of cumulative cases were: Brazil (584,016 cases), Peru (178,914 cases), and Chile (113,628) cases). But when adjusting by cases by 10^6 population, the countries with the largest cumulative cases were Chile (6,066·99 cases × 10^6 population), Peru (5,592·94 cases × 10^6 population), and Panama (3,497·59 cases × 10^6 population). On the other hand, Mexico, Ecuador and Brazil were the countries with the highest CFR with 11·58%, 7·42%, and 5·57%, respectively (Table 1).

**Table 1.** Epidemiological summary of COVID-19 epidemiology in Latin American countries up to June 4^th^ 2020.

| Country | Reported Cases | Cases x 10<sup>6</sup> pop* | Deaths | CFR | Days after introduction | Re | CI95% | Reported Cases<sup>50</sup> | Cases<sup>50</sup> x 10<sup>6</sup> pop | Re<sup>50</sup> | CI95% | Genome Sequences Included |
| --- | --- | --- | --- | --- | --- | --- | --- | --- | --- | --- | --- | --- |
| Venezuela | 1952 | 67.61 | 20 | 1.02 | 82 | 1.44 | 1.35 to 1.53 | 345 | 11.95 | 1.06 | 0.91 to 1.24 | 0 |
| Costa Rica | 1157 | 231.43 | 10 | 0.86 | 89 | 1.11 | 1.02 to 1.21 | 693 | 138.62 | 1.06 | 0.95 to 1.17 | 5 |
| Uruguay | 828 | 240.05 | 23 | 2.78 | 82 | 1.02 | 0.93 to 1.12 | 652 | 189.02 | 1.08 | 0.96 to 1.20 | 7 |
| Nicaragua | 1118 | 172.92 | 46 | 4.11 | 78 | 1.68 | 1.55 to 1.82 | 16 | 2.47 | 1.1 | 0.49 to 2.08 | 0 |
| Paraguay | 1070 | 153.82 | 11 | 1.03 | 85 | 1.12 | 1.02 to 1.22 | 249 | 35.80 | 1.14 | 0.94 to 1.36 | 0 |
| Cuba | 2107 | 185.83 | 83 | 3.94 | 82 | 1.04 | 0.98 to 1.11 | 1611 | 142.09 | 1.2 | 1.11 to 1.28 | 0 |
| Panama | 14609 | 3497.59 | 357 | 2.44 | 87 | 1.18 | 1.15 to 1.21 | 6021 | 1441.51 | 1.24 | 1.20 to 1.29 | 1 |
| Argentina | 19255 | 432.75 | 583 | 3.03 | 90 | 1.31 | 1.29 to 1.34 | 3423 | 76.93 | 1.25 | 1.19 to 1.32 | 24 |
| Chile | 113628 | 6066.90 | 1275 | 1.12 | 92 | 1.31 | 1.30 to 1.32 | 11296 | 603.12 | 1.28 | 1.25 to 1.32 | 147 |
| Ecuador | 46998 | 2750.94 | 3486 | 7.42 | 91 | 1.06 | 1.04 to 1.07 | 11183 | 654.58 | 1.31 | 1.27 to 1.35 | 0 |
| Honduras | 5690 | 593.48 | 234 | 4.11 | 83 | 1.23 | 1.18 to 1.28 | 899 | 93.77 | 1.32 | 1.19 to 1.46 | 0 |
| Colombia | 33354 | 671.80 | 1045 | 3.13 | 86 | 1.31 | 1.29 to 1.33 | 5949 | 119.82 | 1.33 | 1.28 to 1.39 | 68 |
| Guatemala | 5760 | 333.96 | 143 | 2.48 | 82 | 1.32 | 1.27 to 1.36 | 688 | 39.89 | 1.39 | 1.25 to 1.54 | 0 |
| Bolivia | 11638 | 1025.09 | 400 | 3.44 | 84 | 1.37 | 1.34 to 1.41 | 1167 | 102.79 | 1.43 | 1.31 to 1.55 | 0 |
| Dominican Republic | 18040 | 1697.54 | 516 | 2.86 | 86 | 1.15 | 1.12 to 1.17 | 6416 | 603.74 | 1.45 | 1.39 to 1.51 | 0 |
| Peru | 178914 | 5592.94 | 4894 | 2.74 | 89 | 1.22 | 1.21 to 1.23 | 25331 | 791.86 | 1.47 | 1.44 to 1.50 | 1 |
| Brazil | 584016 | 2788.07 | 32548 | 5.57 | 100 | 1.3 | 1.30 to 1.31 | 25262 | 120.60 | 1.53 | 1.51 to 1.56 | 84 |
| Mexico | 101238 | 802.26 | 11728 | 11.58 | 89 | 1.28 | 1.28 to 1.30 | 13842 | 109.69 | 1.57 | 1.52 to 1.62 | 20 |
| El Salvador | 2705 | 421.29 | 51 | 1.89 | 78 | 1.2 | 1.14 to 1.27 | 695 | 108.24 | 1.59 | 1.43 to 1.76 | 0 |

\*Demographic information was obtained as reported by the European Centre for Disease Prevention and Control, <sup>50</sup>Correspond to data estimated until the day 50 of SARS-CoV-2 introduction to each country. Effective Reproductive Numbers (Re) were estimated using Maximum Likelihood.

The overall Re for LATAM estimated through ML was 1·27 (CI95% 1·26 to 1·27). The epidemic curve and time dependent analysis is showed in Figure 1. The portion of the epidemic curve that best fitted exponential growth was estimated between 03-19-2020 and 05-31-2020, with a growth rate of 0·05%, and Re of 1·342 (IC95% 1·341 to 1·342) during that period of time. Sensitivity analysis showed that Re remained within the 95%IC only for a small choice of time windows, and that varied significantly across different GT distributions (data not shown). In order to compare the evolution of the pandemic in each LATAM country over time, we analyzed the number of cumulative cases per 10^6 population at day 50 of introduction to each country. At this time, the country with the highest number of cumulative cases per 10^6 population was Panama, followed by Peru, and Ecuador (Table 1), although the highest Re corresponded to Salvador, Mexico and Brazil.

**Figure 1.**
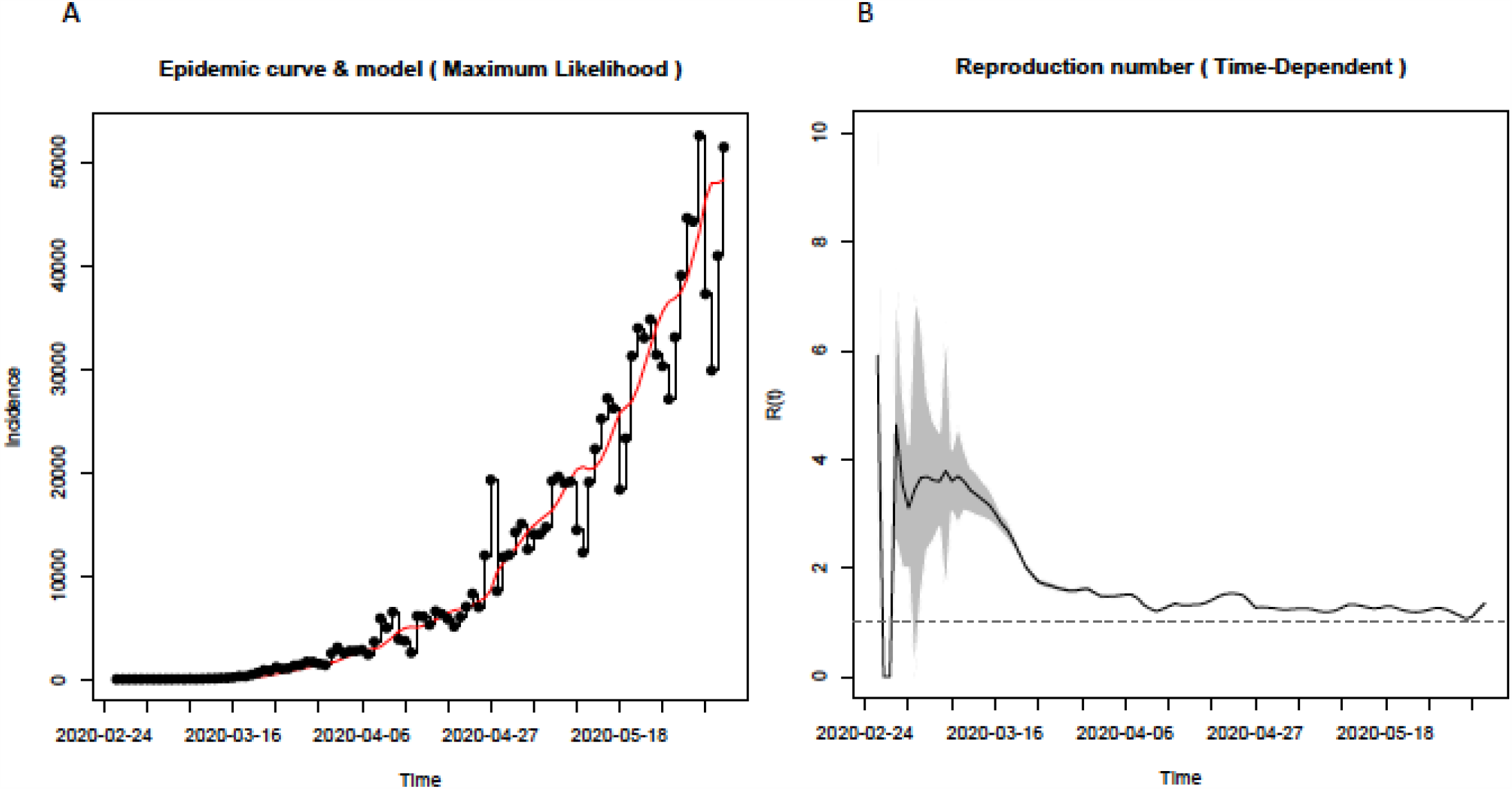
Epidemic curve estimated through Maximum Likelihood (A), and Time-dependent analysis of Effective Reproductive Number, Re (B) in Latin America up to June 4^th^, 2020.

### Phylogenetic Analysis of SARS-CoV-2 in LATAM

A total of 439 genomic sequences were retrieved from GISAID library,^11^ and after quality clearance 357 genomic sequences, from 8 countries, were finally included for analysis (Supplementary Table 1). Substitution rates for every data set is showed in Supplementary Table 2. The PHI test did not find statistically significant evidence for recombination for any of the tested data set. The genomic phylogenetic relation by both, ML and PANGOLIN, showed that SARS-CoV-2 in LATAM includes at least 18 different lineages, with a predominance of B.1 and B.1.1 lineages all across the continent representing 66% of the total included sequences (Supplementary Table 3, Figure 2). Early sequences included the most ancestral A and B lineages that started the pandemic in Hubei, China, indicating the premature SARS-CoV-2 circulation in LATAM through travelers. However, there were important differences at country level. Although lineage B.1 were highly reported across different countries; in some other like Mexico and Uruguay, lineages A.1 and A.5 predominated (Supplementary Figure 1).

**Figure 2.**
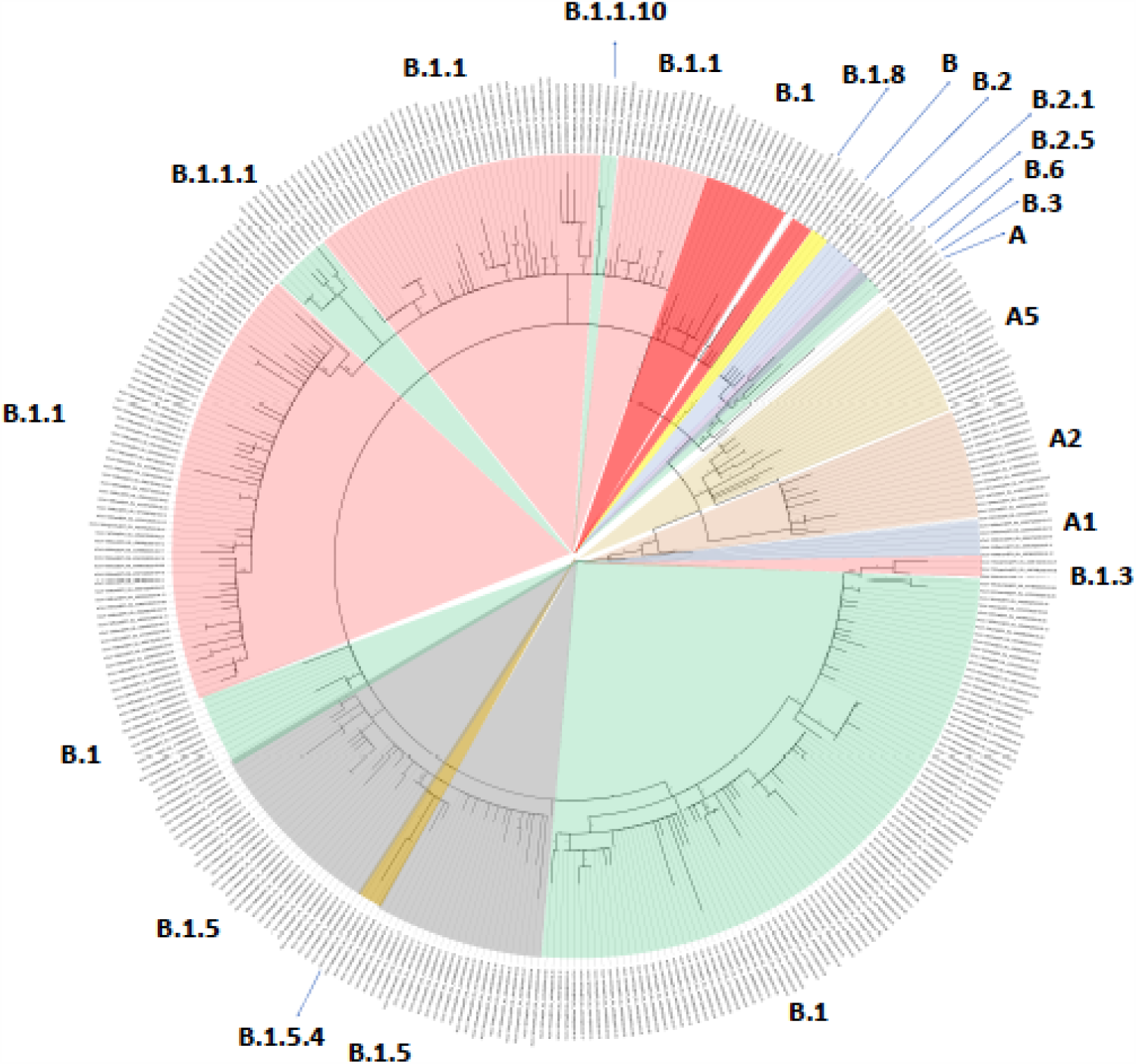
Maximum likelihood phylogenetic relationships between SARS-CoV-2 full genome sequences of Latin America data set. Branch lengths estimated as nucleotide substitutions per site.

### Coalescent and Phylodynamic Analysis

To properly contrast observed epidemiologic curves with phylodynamic inferences, we performed subgroup analysis including only countries with at least five reported genomic sequences and estimating epidemiological observed Re up to the date of the last sampled sequence (LSS). Therefore, only seven countries were finally included in the subgroup analysis; Argentina (LSS 04-19-2020, n = 24), Brazil (LSS 04-27-2020, n = 84), Colombia (LSS 04-07-2020, n = 68), Mexico (LSS 03-21-2020, n = 20), Chile (LSS 04-06-2020, n = 147) Costa Rica (LSS 03-25-2020, n = 5) and Uruguay (LSS 04-21-2020, n = 7). Posterior summary of epidemiological and evolutionary and demographic parameters for each dataset are shown in Supplementary Table 4.

After demographic model estimation, phylodynamic analysis showed an underestimation of the daily incident cases in a factor that varied among countries between 2 to 2200 times compared with data reported to surveillance systems (Figure 3, Table 2). According to our phylodynamic estimations the percentage population infected up to the date of LSS for each country were for Argentina 0·38% (IC95% 0·33 to 0·61), Brazil 1·54% (IC95% 0·86 to 3·37), Chile 1·18% (IC95% 0·76% to 2·13), Colombia 0·22% (0·15 to 0·41), Costa Rica 7·81% (IC95% 5·11 to 14·57), Mexico 0·10% (0·09 to 0·19), and Uruguay 3·5% (2·16 to 8·45), and the percentage of infections diagnosed were for Argentina 4·32 (IC95% 2·71 to 4·94), Brazil 62·16% (IC95% 28·36 to 100), Chile 2·03% (1·12 to 3·13), Colombia 2·15% (1·43 to 0·78), Costa Rica 0·05% (0·02 to 0·07), Mexico 0·17% (IC95% 0·09 to 0·19), Uruguay 0·44 IC95% (0·18 to 0·72). However, when contrasting epidemiological and phylodynamic inferred Re, we did not find statistically significant differences at the cut-off time, except for Chile and Mexico, Table 2. Finally, temporal analysis of the introduction of SARS-CoV-2 revealed that the virus was circulating at least during 21 days before viral detection, except for Uruguay, in which the period was 10 days, and Costa Rica, in which there were not differences between the time of first case notification and TMRA.

**Figure 3.**
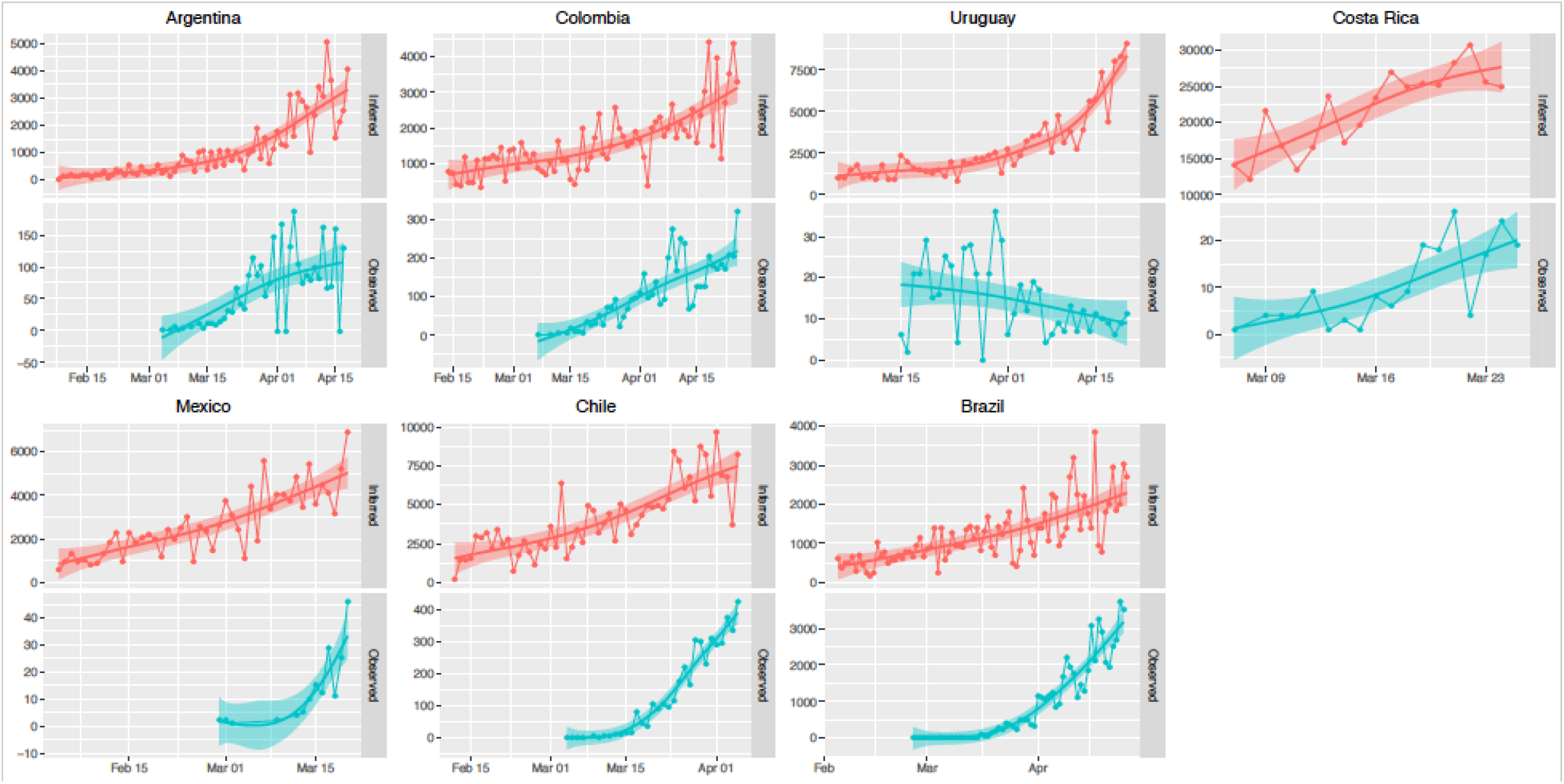
Contrast of inferred (red) and observed (blue) daily incident cases by country since the time to most recent ancestor (TMRA) to last sampled date (LSD). Lines were fitted using a gamma model. Shaded areas correspond to the IC95%. Please note that the scale varies for each country.

**Table 2.**
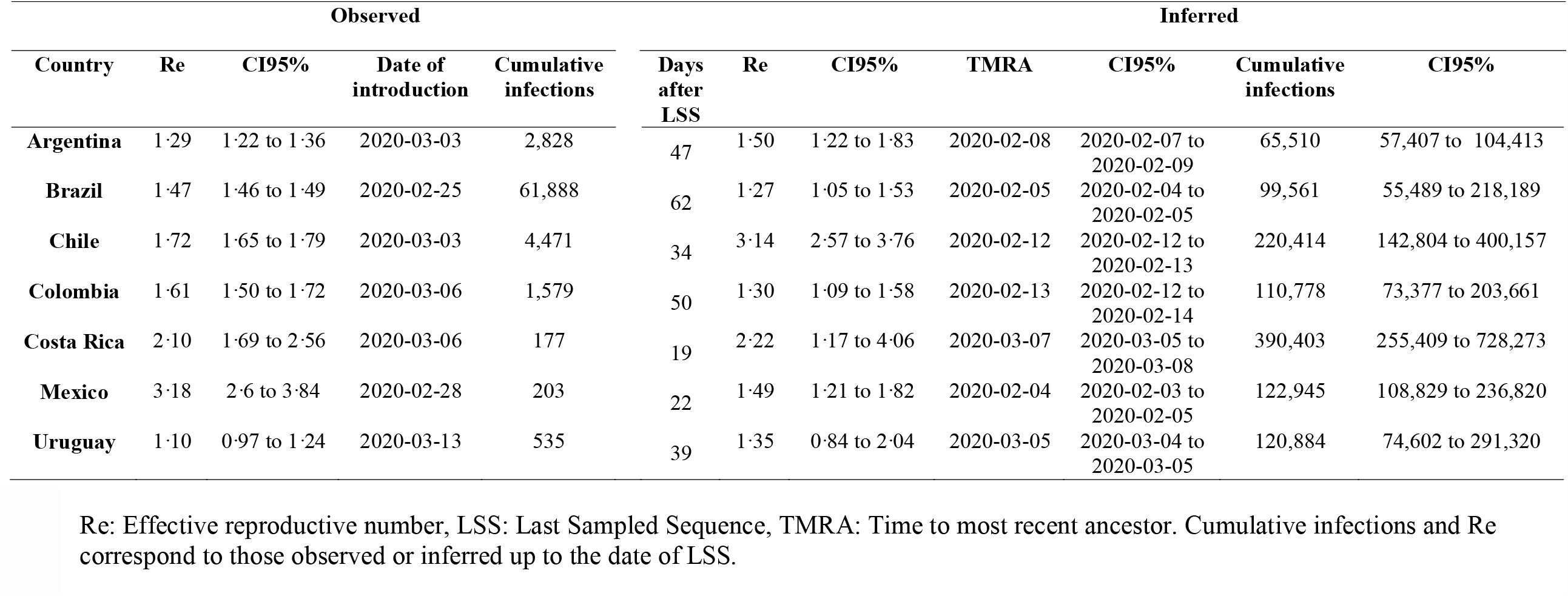
Contrast of epidemiologic and phylodynamic data in Latin America up to the date of last sampled sequence reported for each country.

| Observed |  |  |  |  | Inferred |  |  |  |  |  |  |
| --- | --- | --- | --- | --- | --- | --- | --- | --- | --- | --- | --- |
| Country | Re | CI95% | Date of introduction | Cumulative infections | Days after LSS | Re | CI95% | TMRA | CI95% | Cumulative infections | CI95% |
| Argentina | 1.29 | 1.22 to 1.36 | 2020-03-03 | 2,828 | 47 | 1.50 | 1.22 to 1.83 | 2020-02-08 | 2020-02-07 to 2020-02-09 | 65,510 | 57,407 to 104,413 |
| Brazil | 1.47 | 1.46 to 1.49 | 2020-02-25 | 61,888 | 62 | 1.27 | 1.05 to 1.53 | 2020-02-05 | 2020-02-04 to 2020-02-05 | 99,561 | 55,489 to 218,189 |
| Chile | 1.72 | 1.65 to 1.79 | 2020-03-03 | 4,471 | 34 | 3.14 | 2.57 to 3.76 | 2020-02-12 | 2020-02-12 to 2020-02-13 | 220,414 | 142,804 to 400,157 |
| Colombia | 1.61 | 1.50 to 1.72 | 2020-03-06 | 1,579 | 50 | 1.30 | 1.09 to 1.58 | 2020-02-13 | 2020-02-12 to 2020-02-14 | 110,778 | 73,377 to 203,661 |
| Costa Rica | 2.10 | 1.69 to 2.56 | 2020-03-06 | 177 | 19 | 2.22 | 1.17 to 4.06 | 2020-03-07 | 2020-03-05 to 2020-03-08 | 390,403 | 255,409 to 728,273 |
| Mexico | 3.18 | 2.6 to 3.84 | 2020-02-28 | 203 | 22 | 1.49 | 1.21 to 1.82 | 2020-02-04 | 2020-02-03 to 2020-02-05 | 122,945 | 108,829 to 236,820 |
| Uruguay | 1.10 | 0.97 to 1.24 | 2020-03-13 | 535 | 39 | 1.35 | 0.84 to 2.04 | 2020-03-05 | 2020-03-04 to 2020-03-05 | 120,884 | 74,602 to 291,320 |

Re: Effective reproductive number, LSS: Last Sampled Sequence, TMRA: Time to most recent ancestor. Cumulative infections and Re correspond to those observed or inferred up to the date of LSS.

## Discussion

SARS-CoV-2 transmissibility and epidemiology have varied widely across the world,^18^ apparently related with effectiveness of containment measures, and the time of their implementation. After between the first 100 days of introduction, LATAM have become the epicenter of the COVID-19 pandemic. The magnitude it can reach have raised concern considering the complex sociopolitical scenario in the region^19^ and also the differential impact that the pandemic can have, not only in terms of burden of disease directly derived from COVID-19, but also from diverting public health efforts to only one disease, and from economic effects of containment measures.^5^

To assess the real magnitude of the pandemic will help to estimate the real role and need of containment measures, and will help to propose strategies for adapting them to the current needs, helping balance cost-benefit of those measures. At this respect, our results revealed an overall agreement in terms of transmissibility between epidemiological observed and phylodynamic inferred data, but suggested an overall under detection of COVID-19 cases in LATAM. Importantly, the country with the largest inferred number of underreported cases was Costa Rica, one of the countries with lower cases detected through traditional epidemiological surveillance, while countries with largest cumulative cases like Brazil, Chile and Argentina had a lower estimated underreporting.

Notwithstanding, this data should be interpreted cautiously. In the case of Costa Rica only samples collected very early in the pandemic were included. This implies that the increased variability detected could be product of multiple introductions into the country at the beginning of the pandemic rather than from cumulated mutations through transmission chains in time, which could lead to an overestimation of the number of cases. This explanation is supported by the fact that phylogenetic analysis revealed different linages circulating early in the pandemic, and that the TMRA coincided with the time of epidemiological detection of the virus in the country, showing that SARS-CoV-2 was circulating in Costa Rica for less than a week before first case detection. This could be also the case of Uruguay, in which only seven sequences were finally included for phylodynamic analysis. In this country, after SARS-CoV-2 detection, public health authorities lead to early declaration of the sanitary emergency, with high number of tests per millions of habitants compared with neighbor countries like Brazil, Argentina and Paraguay. In this particular country, availability of more recent genome sequences would be desirable in order to assess the effect of the implemented measures.

Additionally, more than the 50% of the included sequences for Brazil and Mexico were sampled in a same geographical area. This could entail that cumulated variability were product of local transmission chains more than national level transmission, leading to an underestimation of the cases for these countries. Nonetheless, new available data can help to surpass this limitation, particularly for Brazil. According to Candido et al,^20^ when a data set of 427 new sequences were added to the GISAID available sequences up to 30 April, a strong spatial representativity of the country was found. Therefore, a subsequent analysis of this data could provide insights in to the real magnitude of the pandemic in this country.

On the other hand, although our estimations of the percentage of population infected for Argentina, Brazil, Chile and Colombia significantly differ from modelled seroprevalence estimations for those countries,^21^ our estimations of the percentage of undiagnosed infections are similar to those showed by seroprevalence studies. According to our results, at least the 96% of the infections in the analyzed countries (excluding Brazil) are not diagnosed, that agrees with serological community based studies in Hubei, China that suggests that 97% of infections might have gone undiagnosed during the epidemic, ^22^ and in Geneva, Switzerland that showed that for every confirmed case there were 11.6 infections in the community.^23^

Regarding transmissibility, the Re was in good agreement for every country, except for Chile and Mexico, and is consistent with previous estimations for Latin American countries^24-27^. This has two important implications: First, the good agreement at this respect validates previous transmission dynamics of viral shedding estimated by He, X, *et al*,^14^ suggesting that transmission in LATAM have had a similar behavior to the observed in other countries, and second, rise concern about the effectiveness of containment measures implemented in Chile and Mexico. On the other hand, the fact that most of the countries had a detection lag of at least 21 days, highlights the need of improvement of index case detection in local healthcare systems.

As reported by others for the Europe and North American outbreaks,^15,28^ in LATAM the B.1 lineage was one of the first lineages that was stablished and was the most prominent lineage that spread with B.1.1 lineage. Although it has been claimed that B.1 lineage harbors a D614G mutation in the Spike protein that make the virus more contagious,^29^ our results about the early circulation of B.1 lineage in LATAM and the fact that not all the countries that has B.1 has a high prevalence over other lineages, it cannot be confirmed that that D614G showed high fitness as has been reported.^28,30^

Nevertheless, our estimations have some limitations. First, the quality of the inferred phylodynamic data for each country depends on the extent of the geospatial genome sampling. For some countries like for example Brazil and Mexico, samples were limited in a high proportion to a particular geographical area within the countries, and hence estimations may not necessarily reflect data generalizable to the entire country. On the other hand, the limited availability of clinical and epidemiological metadata do not allow to properly evaluate transmission chains within the countries. Therefore, observed variability could not be related to cumulated mutation through transmission chains, but to different introductions to each country, as suggested by the co-circulation of multiple lineages. The lack of availability of enough samples from other countries limited the number of countries to be included in the phylodynamic analysis.

Finally, our results support that epidemiological and genomic surveillance are two complementary approaches for analysis of transmission and epidemiological patterns within and between countries. Overall Re estimation for LATAM through phylodynamic analysis showed good agreement with observed Re through epidemiological surveillance even when a large amount of sequences were not available for every country. The evidence suggests that even with a low number of sequences, proper estimations of Re could be performed. This could provide important insights, especially at the beginning of an epidemic when epidemiological surveillance systems are adjusting follow-up and control measures. At the same time, we found that the lack of clinical and epidemiological metadata for reported sequences difficult making phylodynamic inferences. We suggest that countries, especially developing countries, should consider to add genomic surveillance to their systems for monitoring and adapting epidemiological surveillance of SARS-CoV-2.

## Data Availability

Data will be available upon request

## Authors contributions

Diana M. Rojas-Gallardo: Study design, data analysis, data interpretation, and manuscript writing*, Sandra C. Garzón-Castaño: data analysis, data interpretation, and manuscript writing, Natalia Millán: data collection, data analysis, and manuscript writing, Erika V. Jiménez-Posada: data collection, data analysis, and manuscript writing, Marlen Martínez-Gutiérrez: data analysis, data interpretation and manuscript writing, Julian Ruiz-Saenz: data analysis, data interpretation, data analysis, and manuscript writing, Jaime A. Cardona-Ospina: Study design, data analysis, data interpretation, and manuscript writing*

*These two authors contributed equally.

## Acknowledgements

We would like to acknowledge Grupo de Investigación SIRIUS, Facultad de Ingenierias, Universidad Tecnológica de Pereira, for providing resources for performing high performance computational analysis.

## Declaration of interests

The authors have nothing to disclose.

## Funding

None.

## Notes

### Competing Interest Statement

The authors have declared no competing interest.

### Funding Statement

No external funding has been received

### Author Declarations

This is a study based on epidemiological data and does not require IRB approval

